# Multimorbidity increases susceptibility to myocardial injury via dysregulated cardiac macrophage activation and the development of a cardiomyopathy phenotype

**DOI:** 10.1101/2024.04.05.24305379

**Authors:** Florence Y Lai, Natasha Beasley, Shameem Ladak, Charles Solomon, Adewale S Adebayo, Sophia Sheikh, Marius Roman, Lathishia Joel-David, Hardeep Aujla, Tom Chad, Kristina Tomkova, Gianluigi Condorelli, Mustafa Zakkar, Marcin J Woźniak, Tom Webb, Veryan Codd, Gavin J Murphy

## Abstract

**Background:** This study investigated mechanisms underlying the association between multimorbidity (MM) and increased susceptibility to myocardial injury and organ failure following cardiac surgery.

**Methods:** K-means clustering was performed using data from five cardiac surgery cohorts with high prevalence of MM. The resulting Clusters were explored using single nuclei RNA sequencing (snRNAseq) of atrial biopsies collected at surgery from one of the study cohorts. Mechanisms were validated using causal inference methods in UK Biobank and aged cardiomyocytes exposed to ischaemia reperfusion injury (IRI) *in vitro*.

**Results:** K-means clustering using pre-surgery biomarkers of haematopoietic, cardiac, metabolic, liver, and renal disease identified two MM clusters. Cluster 1, characterised by higher baseline troponin I and interleukin-6 values, iron deficiency, anaemia, and immunological ageing, demonstrated significantly higher rates of myocardial injury (66% versus 52%) and multiple organ dysfunction (81% versus 57%) relative to Cluster 2.

snRNAseq data from Cluster 1 demonstrated inflammageing characterised by enrichment for cardiomyopathy networks in cardiomyocytes, NF-kB and IL2 in monocyte derived macrophages (MDM), and pro-fibrotic and redox inflammation signaling in tissue resident macrophages (TRM). Cluster 2 showed enrichment for translation, type 1 inflammation, and immune activation in cardiomyocytes, and acute-phase immune responses in MDM and TRM.

In UK Biobank, genetic modification of genes differentially expressed between clusters altered 90-day mortality post-surgery. Gene silencing of key regulatory nodes enriched in Cluster 1 cardiomyocytes including PDE1c (Ca^2+^homeostasis) and SNAI1 (TGFβ-SMAD), attenuated cardiomyocyte de-differentiation and susceptibility to IRI *in vitro*.

**Conclusions:** Inflammageing associated cardiomyocyte dedifferentiation represents a target for myocardial protection in people with MM.

**Study Schematic:** MaRACAS, the Observational Case Control Study to Identify the Role of MV and MV Derived Micro-RNA in Post CArdiac Surgery AKI, REVAKI-2, Revatio® for the Prevention of AKI -2 trial, REDWASH, Red Cell washing for the Prevention of Organ Injury after Cardiac Surgery Trial, OBCARD, the Case Control Study to Identify the Role of Epigenetic Regulation of Genes Responsible for Energy Metabolism and Mitochondrial Function in the Obesity Paradox in Cardiac Surgery, COPTIC, the Coagulation and Platelet Laboratory Testing in Cardiac Surgery study.

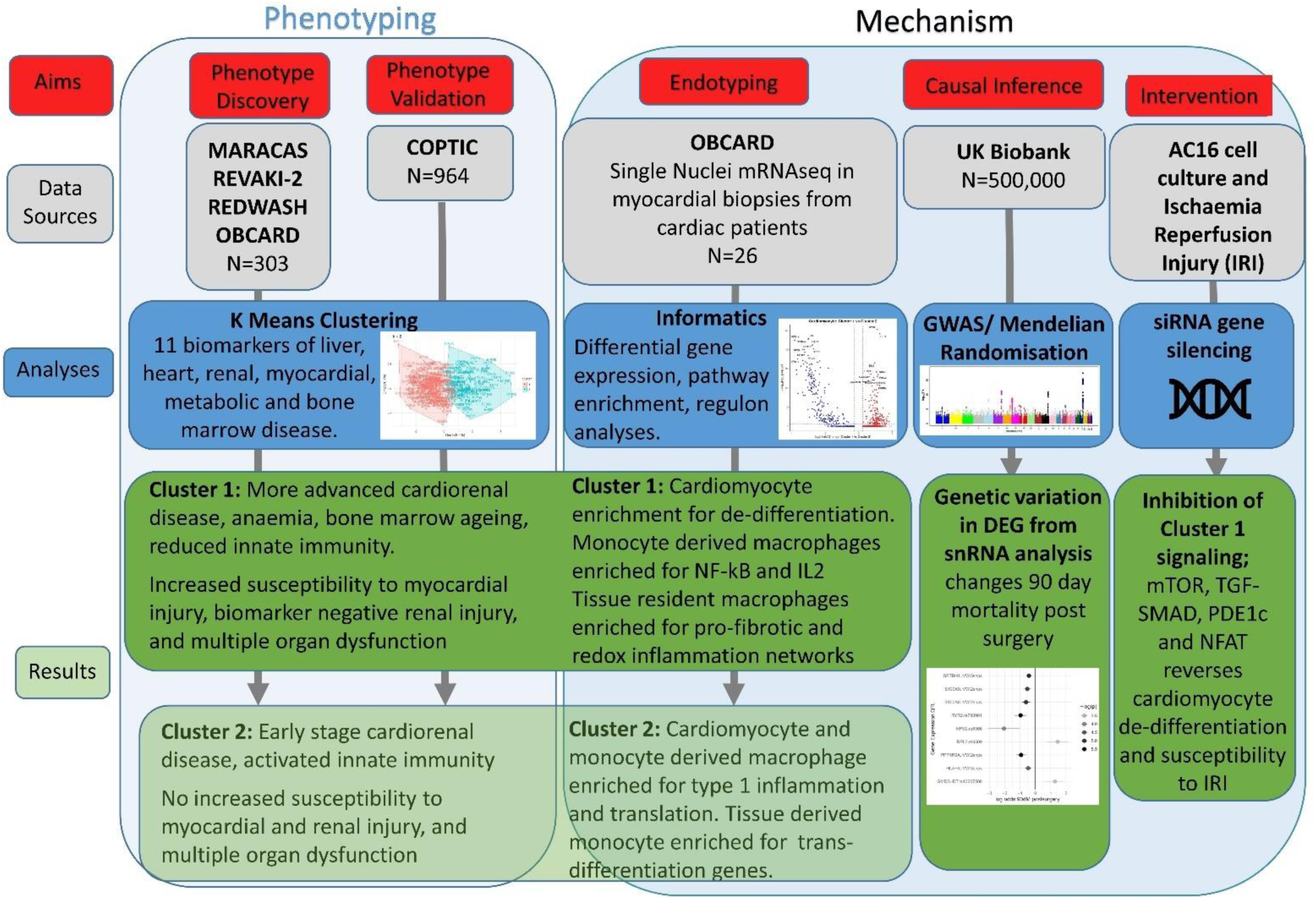

## Introduction

The long-term benefits of cardiac surgery are offset by early organ injury affecting the heart, lungs, brain, and kidneys. These complicate >30% of procedures where they are the primary cause of early death or result in delayed recovery, lower quality of life, and increased healthcare resource use. Despite decades of research, there are no clinically effective organ protection interventions.(1–4)

People referred for cardiac surgery increasingly present with Multimorbidity (MM), currently defined as the presence of two or more long-term health conditions.(5, 6) This population demonstrates increased susceptibility to organ injury and death relative to people without MM.(7) We hypothesised that disease processes associated with MM may offer novel targets for pre-surgery organ protection.

MM demonstrates substantial heterogeneity of the type and severity of underlying chronic diseases.(8) We addressed this by using quantitative biomarkers of cardiac, metabolic, renal, liver, and haematologic disease severity from 5 intensively phenotyped cardiac surgery cohorts (9–13) where recruitment used risk scores (2, 14) to ensure MM enrichment. We applied unbiased clustering analyses to these data to identify different MM phenotypes and assess their susceptibility to organ injury. Then, using single nuclei RNA sequencing (snRNAseq) of myocardial biopsies from one of the study cohorts (12) we investigated the differences in the transcriptome between Clusters. Finally, we evaluated the causality of mechanisms associated with susceptibility to injury identified from the transcriptome analyses using causal inference analyses (Mendelian randomisation) in UK Biobank and *in vitro* experiments.

## Methods

### Study ethics

UK Human Research Authority ethics approvals were 18/WS/0148, 21/NW/0157 and 21/SC/0118. The clustering analyses were pre-specified.(15) All the participants had provided consent for secondary research using their data. The results are reported as per the STrengthening the Reporting of Observational Studies in Epidemiology (STROBE) statement.(16)

### Data Sources

The primary clustering analysis included data from 277 participants of three studies (9, 10, 13) that recruited cohorts enriched for MM (AKI risk score>18 (2) or LVBT Score>22 (14)) between 2013 and 2020 at 3 UK centres. In addition, 26 participants from a fourth study (ObCARD, recruited 2021-2024) (17) who demonstrated MM enrichment and had single nuclei RNA sequencing (snRNAseq) data available from right atrial biopsies collected at surgery, were included. All 4 studies used common outcome definitions, had low levels of missing data, and performed biomarker and flow cytometry analyses in a single laboratory using standardised methods.(8) The primary analysis cohort therefore included 303 study participants with complete baseline biomarker data.

To externally validate the results of the primary analysis, K-means clustering was also performed using the same baseline biomarkers in participants enrolled in the Coagulation and platelet laboratory testing in cardiac surgery (COPTIC) study at a 4^th^ UK centre.(11) Included participants met the MM enrichment criteria (AKI risk score>18), and had complete baseline biomarker data.

Causal inference analyses used Imputed Genotype Data v3 from UK Biobank (Application 77596).

### Input variables for cluster analysis

The clustering analyses used continuously distributed baseline patient characteristics and pre-surgery serum biomarkers of long-term conditions common to all 5 studies. These were: Age, body mass index (BMI), serum bilirubin, estimated Glomerular filtration rate (eGFR) from serum creatinine, platelet count, haemoglobin concentration (Hb), serum transferrin saturation, ferritin, interleukins (IL)-6 , IL-8 and high sensitivity serum Troponin I (Enzo® Troponin I (human) ELISA kit, Enzo Life Sciences, Ann Arbor, MI, USA).

### Outcomes

Clinical outcomes were restricted to those measured prospectively in the individual studies. **For the primary analysis,** myocardial injury was defined by peak serum hsTroponin I within 24 hours postoperatively as defined by Devereaux and colleagues, (18) post-operative acute kidney injury (AKI) was defined using KDIGO criteria, (19) urinary Neutrophil Gelatinase Associated Lipocalcin (NGAL, BioPorto, Hellerup, Denmark) and tissue inhibitor of metalloproteinases-2 *insulin-like growth factor-binding protein 7 (TIMP2*IGFBP7 EIA kit, EKF diagnostics, Cardiff, UK). Acute lung injury was determined using Berlin criteria.(20) **In the validation cohort**, the main clinical outcome was in-hospital death or major adverse cardiovascular events (MACE) defined as KDIGO defined Stage 3 acute kidney injury, myocardial injury, and stroke. Myocardial injury was defined as suspected MI or new low cardiac output syndrome as defined in the primary publication.(11)

Assessments of platelet and leucocyte activation at baseline common to three studies (9, 10, 13) in the primary analysis cohort included flow cytometry (Cyan ADP; Beckman Coulter) in whole blood for P selectin/CD62P (Abcam, Cambridge, UK), PE-coupled CD41 (Affymetrix, Santa Clara, CA, USA) and leucocyte activation (CD64, CD163; Affymetrix). Microvesicle concentrations were calibrated using the NanoSight NS500 (Malvern Pananalytical), with surface markers/ cell derivation identified using flow cytometry as described previously.(10) Validation of the NanoSight, FACS data and Multiplate data was as per the primary study publications.(9, 10, 13) Baseline platelet function was assessed using the Multiplate® analyser (Roche, Rotkreuz, Switzerland). In the validation cohort, haematopoiesis markers were measured on the Sysmex XN-1000 (Milton Keynes, UK) and plasma levels of immune ageing biomarkers; CCL11/ Eotaxin, CX3CL1 /Fractalkine, GDF-15, IL5, CXCL9/MIG, CCL3/MIP-1 alpha, high sensitivity C Reactive Protein, and Serpin were measured at baseline using Luminex® Multiplex Assays (Luminex, Genk, Belgium) on the Magpix® platform (Luminex).

### Clustering analysis of clinical data

We applied k-means clustering to the primary and validation cohorts independently using the R-package *cluster*. Input variables were log-transformed if skewed. Pearson’s correlation coefficient was calculated to identify any highly correlated variables. Variables that are highly correlated (Pearson’s correlation >0.6) were filtered. Missing data in input variables were low (range 0% - 8.5%). To maximise the inclusion of patient data, missing data were imputed using random forest imputation (R package *missForest*). Variables were scaled to the standard normal distribution with a mean of 0 and a standard deviation of 1 prior to clustering. The optimal number of clusters was determined based on Silhouette distance. We additionally considered the NbClust program in R (21) and used the majority rule of the 23 indices of cluster cohesiveness and separation to confirm the optimal cluster numbers.

After the clusters were identified, we characterised the clinical phenotypes based on participants’ baseline characteristics and their post-op organ injury outcomes. Differences in characteristics across phenotypes were compared using the Wilcoxon test for continuous variables and the chi-square test for categorical variables.

### single nuclei RNA sequencing in myocardial biopsies

snRNAseq data was available for 26 participants from the OBCARD study (17) who were included in the primary analysis cohort cluster assignment. snRNAseq was performed as described for single-nucleus isolation from human atrial biopsies (30 – 100 mg) collected prior to cardiopulmonary bypass from the right atrial appendage, (22) immediately snap-frozen in liquid nitrogen, and stored at -80⁰C. After nuclei isolation, cDNA amplification and library construction were performed using Chromium Single Cell 3’ Gel-bead in Emulsion (GEM) and Chromium Single Cell 3’ Library kits v3.1. (#1000128, 10x Genomics, Pleasanton, United States) following the manufacturer’s instructions.(23) The resulting cDNA was quality-checked using the Agilent Bioanalyzer High Sensitivity DNA kit (Agilent Technologies, Santa Clara, United States). Libraries were constructed, including fragmentation, end-repair, A-tailing, and adaptor ligation steps. A unique sample index was added to each sample.

Following library construction, samples were quality-checked, and libraries were purified and eluted using Buffer EB (#19086, Qiagen, Hilden, Germany), and SPRIselect beads (#B23317, Beckman Coulter, Pasadena, United States). Samples were sequenced paired-end on Illumina NovoSeq at 2.5×108 reads per sample and analysed using existing pipelines as described below. Reads in fastq formats were QC’ed and processed using 10X Genomics CellRanger v6.0.1.

### snRNA-seq data processing and quality control

Raw gene expression matrices were imported into R (v4.3.1) using the Seurat package (v5.3.1) via the Read10x function (24, 25). Nuclei with >5% mitochondrial gene content (percent.mt), calculated based on genes annotated with the “MT-“ prefix were excluded, to remove low quality or apoptotic nuclei. Additionally, nuclei with <200 and >12500 detected genes were removed. Potential doublets were identified and removed using DoubletFinder (v2.0.6) by optimizing pK via parameter sweeps (BCmetric) and estimating expected doublets at ∼1% of nuclei adjusted for homotypic doublet portion (26).

### Data normalisation and integration

Each dataset was independently normalised (NormalizeData), variable features were identified (FindVariableFeatures using 2,000 features), scaled (ScaleData) and principal component analysis was performed (RunPCA, 30 PCs). Datasets were integrated using Seurats RPCA framework and the integrated expression values were computed with IntegrateData (k.weight = 5).

### Cell type annotation and ambient RNA correction

Cell type annotation was performed using a reference-based label transfer approach. A heart reference atlas was generated by downloading a pre-annotated single nuclei dataset by downloading from the Broad Institute Single Cell Portal (SCP1849; ICM_scportal_05.24.2022.h5ad) (27). The reference dataset was converted into a Seurat object and pre-processed using SCTransform, with the SCT assay used for downstream dimensionality reduction (28). PCA was performed on the reference, and anchors between the reference and the query datasets were identified using FindTransferAnchors. Reference-derived cell type labels were transferred to all nuclei using MapQuery. Ambient RNA contamination was removed using DecontX (celda v1.18.2) and the decontaminated counts were used for downstream expression-based analysis.

Macrophages were further subcategorised as tissue-resident (TR) or monocyte-derived (MD) using predefined marker gene sets and Seurat AddModuleScore, with a margin of 0.05, with intermediate cells labelled as ambiguous.

### Differential Expression analysis

Prior to differential expression analysis, cells were down sampled in the larger group (set.seed = 123), controlling for differences in cell numbers between groups. Differential expression analysis was performed using Seurats FindMarkers with a logistic regression model. Genes expressed in >10% of cells in either group with a minimum log2 fold change threshold of 0.25 were tested, and significance was defined as an adjusted p value < 0.05 (Benjamini-Hochberg correction) (29).

### Pathway analysis

Functional enrichment analysis was performed using the ClusterProfiler Bioconductor package (30). Differentially expressed genes were filtered by adjusted p-value (<0.05) and log2 fold change (>0.25), with up- and down-regulated genes analysed separately. Gene symbols were converted to Entrez Gene identifiers using the org.Hs.eg.db (v3.18.0) annotation (31). Gene Ontology (GO) biological process (BP) and KEGG pathway enrichment analysis were conducted using the enrichGO and enrichKEGG functions respectively, with Benjamini-Hochberg correction. Terms with q-values < 0.05 were considered significantly enriched.

For the macrophage analysis, pathway activity inference was performed using PROGENy (v1.24.0) implemented in the decoupleR (v2.8.0) framework (32, 33). Normalised expression values were used as input, and pathway activities were inferred at the single-cell level using the human PROGENy model and the multilayer linear model (run_mlm). Mean pathway activity scores were calculated for each cluster, and relative pathway enrichment was determined based on differences in average pathway activity between groups.

### Regulon analysis

Transcription factor (TF) activity was inferred using the DoRothEA regulon (v1.14.1) and VIPER (v1.36.0) algorithm, implemented in decoupleR (34). TF activity scores were computed for each cell based on the expression of regulon target genes. Differences in TF activity between clusters were assessed using Wilcoxon rank-sum tests, with P-values adjusted using the Benjamini-Hochberg method (FDR < 0.05). Regulons corresponding to significantly differentially active TFs were further examined. To identify functionally supported regulatory interactions, regulon target genes were intersected with differentially expressed genes. Pathway enrichment analysis of regulon target genes was performed using MSigDB Hallmark gene sets (v25.1.1) (35).

### Causal Inference analysis

UK Biobank was used to infer the likely causality of the observed differences in gene expression between clusters and clinical outcomes. SNPs identified as expression quantitative trait loci (eQTLs) were used as instruments for causal inference analysis. Effect sizes of SNP on expression (summary statistics) were obtained from MRbase (36, 37) which were derived from analysis on healthy tissue. Imputed genotype data v3 was obtained from UK Biobank. Per chromosome bgen files were converted and combined to genotype matrix using qctool (38), VariantAnnotation (39) and snpStats (40) packages. Major cardiac surgery procedures were defined using OPCS4 codes (K40–K46, K49, K50, K75). Outcomes were defined as death within 90 days of surgery using the death register. Effect sizes on outcomes were calculated after adjusting for age, sex and 10 principal components. Annotations of UKBiobank SNPs were obtained from NealeLab data (http://www.nealelab.is/uk-biobank) round 2 results. Both summarized data on exposure (gene expression) and outcomes were combined in an MR analysis using the TwoSampleMR package.(37) Where only single instruments were available for the exposure the Wald ratio test was applied. Where multiple instruments were available for exposure, the inverse variance weighted method was applied. The functional annotation of significant genes was obtained from GeneCards (41) and PantherDB.(42, 43)

### In vitro experiments

AC16 human cardiomyocyte cell lines were maintained at 37 °C in 5% CO2 in DMEM: F12 medium (Gibco, 11580546) supplemented with 12.5 % FBS and (44) Simulation of Cluster 1 used 4 hrs exposure to H_2_0_2_ 100µM and IL6 10ng.ml^-1^ to model the ROS and IL-6 signalling observed in *in vivo.* Simulation of Cluster 2 used 4 hrs TNFα 20ng.ml^-1^ and IL-1β 10ng.ml^-1^ exposure to model the Type 1 inflammation (45) observed *in vivo*. Cells cultured only in total medium served as controls. Next Generation bulk mRNAseq (Source Genomic Ltd, Nottingham UK)of AC16 cells was used to estimate homology to the snRNAseq analysis. The combined data was used to propose a model of cardiomyocyte inflammageing analogous to the Cluster 1 phenotype consistent with previously published studies.(46–50)

Gene silencing was achieved by transfection with Lipofectamine RNAiMAX (Invitrogen, Carlsbad, CA, USA, 13778075) transfection reagent and 50 μmol/L final concentration of PDE1C siRNA (ThermoFisher Scientific, Silencer Select; Assay ID: 143972), ERBB3 siRNA (ThermoFisher Scientific, Silencer Select; Assay ID: 146246), SNAI1 siRNA (ThermoFisher Scientific, Silencer Select; Assay ID: 107915) and GATA4 siRNA (ThermoFisher Scientific, Silencer Select; Assay ID: 115249) or Silencer™ Select Negative Control No. 1 siRNA (ThermoFisher Scientific, 4390843) in Opti-MEM reduced serum media (ThermoFisher Scientific, 31985070) according to the manufacturer’s protocol for 24 hours.

Ischaemia reperfusion injury (IRI) was simulated by culture in glucose deficient media in anoxic conditions (95% N_2_ and 5% CO_2_) at 37°C for 4 hrs, followed by replacing glucose free media with total medium at 37°C in a 95% air and 5% CO_2_ (normoxic condition) incubator for 24 hrs. Cells cultured only in normoxic conditions served as controls. Cardiomyocyte injury was assessed using the LDH-Cytox™ Assay Kit (Biolegend, 426401). All experiments used 3-4 repeats.

Total RNA from AC16 cells was isolated using RNeasy kit (Qiagen, Venlo, The Netherlands, 74104) and cDNA was synthesised using Tetro cDNA synthesis kit (Meridian Bioscience, Cincinnati, OH, USA, BIO-65043). Gene expression was quantified by quantitative real time PCR using TaqMan primers (ThermoFisher Scientific, Assay IDs: Hs01005664_m1 and Hs00608023_m1) and SensiFast Probe Hi-ROX kit (Meridian Bioscience, Cincinnati, OH, USA, BIO-82005) on Rotor gene Q (Qiagen) using the manufacturer’s protocol. Relative levels were calculated using the 2−(ΔΔCt) method, and mRNA expression was normalised to housekeeping gene PPIA (ThermoFisher Scientific, Assay ID: Hs99999904_m1).

## Results

### Unsupervised clustering identifies a MM phenotype with increased susceptibility to organ injury

In the primary analysis, unsupervised K-means clustering demonstrated two Clusters (**Figure 1A, sTable 1**). Cluster 1 had a higher proportion of females (29% versus 12%), lower bilirubin, Haemoglobin, Ferritin, Transferrin Saturations, and eGFR, and higher platelet counts, hsTroponin I, IL-6 and NT-proBNP at baseline, versus Cluster 2 (**Table 1, sTable s2**). There were no significant differences between the Clusters for age, or prevalence of diabetes mellitus, chronic kidney disease (eGFR<60 mls.min^-1^.1.73m^-2^), heart failure, angina severity, stroke, liver, lung or neurological disease, history of cancer, or peripheral vascular disease at baseline, type of operation, and duration of cardiopulmonary bypass or aortic cross clamping (**Table 1**).

**Figure 1.**
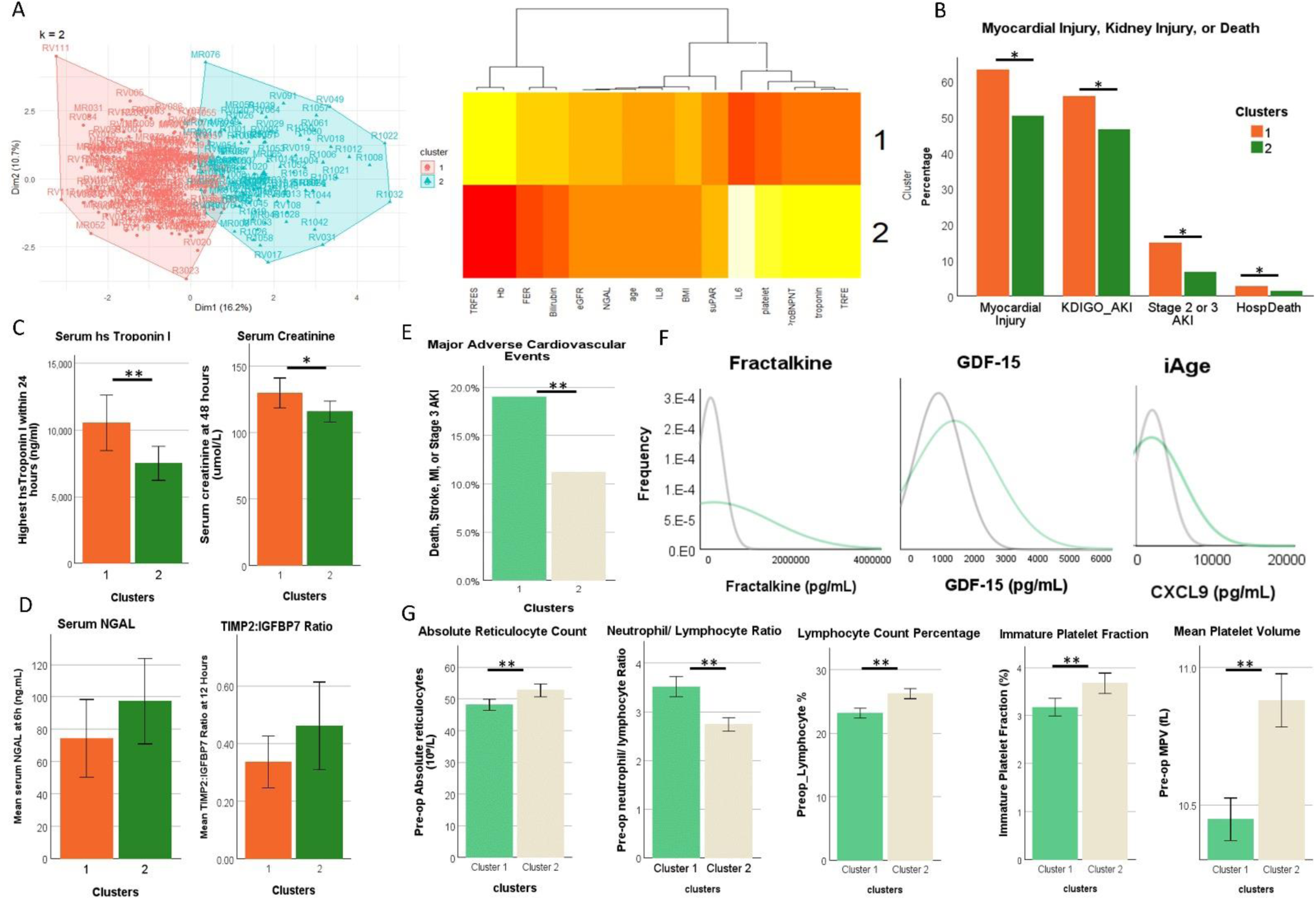
In the primary analysis cohort (n=303) of 4 cardiac surgery studies**; A.** K Means Clustering using 11 MM biomarkers identified 2 clusters. **B.** Cluster 1 was associated with significant increases in Myocardial Injury, Acute Kidney Injury, Multiple Organ Dysfunction and Death versus Cluster 2. **C.** Cluster 1 demonstrated higher serum high sensitivity Troponin I and serum creatinine levels. **D.** Acute Kidney Injury biomarkers were not significantly different between clusters. In the COPTIC validation cohort (n=963), Cluster 1 was associated with; **E.** Significant increases in Death or Major Adverse Cardiovascular Events (MACE), **F.** Elevated biomarkers of immune ageing and, **G.** dysregulated haematopoiesis, myelopoiesis and thrombopoiesis at baseline. *P<0.01 **P<0.001. Statistical tests as per text. GDF-15, Growth Differentiation Factor 15. TIMP2, Tissue inhibitor of metalloproteinases-2. IGFBP7, Insulin-like growth factor-binding protein 7. NGAL, Neutrophil gelatinase-associated lipocalin. CXCL9, Chemokine Ligand 9. AKI, Acute Kidney Injury. KDIGO, Kidney Disease Improving Global Outcomes.

**Table 1.** Baseline clinical and operative characteristics in the Primary analysis Cohort.

|  | All Participants | Cluster 1 | Cluster 2 | p-value |
| --- | --- | --- | --- | --- |
|  | N = 303 | N=122 | N=181 |  |
| **Demographics** |  |  |  |  |
| Age (years) | 74 (68 - 77) | 74.5 (68 - 78) | 73 (68 - 77) | 0.3382 |
| Sex – male | 234 (77%) | 74 (61%) | 160 (88%) | <.0001 |
| BMI (kg.m<sup>-2</sup>) | 30.4 (27.0 - 34.6) | 29.5 (25.4 - 32.4) | 31.0 (27.7 - 34.9) | 0.0037 |
| Ethnicity - Caucasian | 290 (96%) | 111 (91%) | 179 (99%) | 0.0009 |
| **Medical history** |  |  |  |  |
| Canadian Cardiology Society (CCS) Class |  |  |  |  |
| asymptomatic | 105 (35%) | 42 (36%) | 63 (35%) | 0.1504 |
| I | 85 (29%) | 37 (31%) | 48 (27%) |  |
| II | 84 (28%) | 26 (22%) | 58 (32%) |  |
| III & IV | 24 (8%) | 13 (11%) | 11 (6%) |  |
| Obese (BMI >32) | 113 (37%) | 37 (30%) | 76 (42%) | 0.0395 |
| Renal disease | 30 (10%) | 15 (13%) | 15 (8%) | 0.2352 |
| Anaemia (Hb<120 g.dL<sup>-1</sup> F, Hb<130 g.dL<sup>-1</sup> M) | 154 (52%) | 102 (85%) | 52 (29%) | <.0001 |
| Diabetes | 140 (47%) | 60 (50%) | 80 (45%) | 0.3319 |
| Stroke/TIA | 28 (9%) | 10 (8%) | 18 (10%) | 0.6214 |
| Angina (CCS class II, III, IV) /previous MI | 140 (46%) | 52 (43%) | 88 (49%) | 0.3046 |
| Heart failure (NYHA class III, IV) | 64 (21%) | 27 (23%) | 37 (20%) | 0.6152 |
| Hypertension | 11 (4%) | 4 (3%) | 7 (4%) | 1 |
| Hyperlipidemia | 1 (0%) | 0 (0%) | 1 (1%) | 1 |
| Pulmonary hypertension | 58 (19%) | 21 (17%) | 37 (20%) | 0.4835 |
| Extracardiac arteriopathy | 17 (6%) | 9 (8%) | 8 (4%) | 0.2594 |
| Chronic Obstructive Pulmonary Disease | 21 (7%) | 12 (10%) | 9 (5%) | 0.0949 |
| Asthma | 2 (1%) | 0 (0%) | 2 (1%) | 0.5174 |
| Neurological diseases | 5 (2%) | 4 (3%) | 1 (1%) | 0.0847 |
| Cancer | 8 (3%) | 3 (2%) | 5 (3%) | 1 |
| Liver diseases | 3 (1%) | 2 (2%) | 1 (1%) | 0.5671 |
| Arthritis | 2 (1%) | 1 (1%) | 1 (1%) | 1 |
| Multiple (2+ of the above) Long Term Conditions | 245 (81%) | 107 (88%) | 138 (76%) | 0.0129 |
| **Baseline and operative characteristics** |  |  |  |  |
| Haemoglobin concentration (g.dL<sup>-1</sup>) | 128 (114 - 139) | 114 (105 - 123) | 135 (128 - 145) | <0.0001 |
| Haematocrit (%) | 37.9 (34.0 - 40.9) | 34.0 (31.9 - 36.8) | 39.7 (37.2 - 42.7) | <0.0001 |
| Serum creatinine (μmol.l<sup>-1</sup>) | 87 (73 - 105) | 88.5 (70 - 110) | 87 (75 - 103) | 0.9819 |
| PaO2/FiO2 ratio | 457.1 (409.5 - 533.3) | 457 (410 - 533) | 457 (410 - 533) | 0.2724 |
| eGFR (ml/min/1.73 m<sup>2</sup>) | 74 (61 - 93) | 72 (55 - 94) | 76 (64 - 91) | 0.0395 |
| Trial |  |  |  |  |
| MARACAS | 95 (31%) | 22 (18%) | 73 (40%) | <.0001 |
| REDWASH | 56 (18%) | 51 (42%) | 5 (3%) |  |
| REVAKI2 | 126 (42%) | 41 (34%) | 85 (47%) |  |
| OBCARD | 26 (9%) | 8 (7%) | 18 (10%) |  |
| Surgery |  |  |  |  |
| CABG only | 113 (37%) | 38 (31%) | 75 (42%) | 0.1534 |
| Valve only | 92 (30%) | 39 (32%) | 53 (29%) |  |
| both or other cardiac surgery | 97 (32%) | 45 (37%) | 52 (29%) |  |
| Cardiopulmonary Bypass Time (min) | 105 (81 - 130) | 107 (83 - 140) | 103 (81 - 125) | 0.2313 |

Note: Baseline clinical and operative characteristics are summarised as median (Q1-Q3) for continuous variables and n(percentage) for categorical variables.
**Labels:** NYHA New York Heart Association, eGFR, estimated Glomerular Filtration Rate, CABG, Coronary Artery Bypass Grafts, PaO2 Partial pressure of oxygen in arterial blood in mmHg, FiO2, % inspired oxygen concentration as a ratio, BMI, Body Mass Index

Cluster 1 showed increased susceptibility to organ injury post-surgery (**Figure 1B**). including a higher prevalence of myocardial injury (63% versus 50%), Multiple (>2) Organ Dysfunction (MODS, 81% versus 57%), AKI (56% versus 44%), including Stage 2 or 3 AKI (15% versus 7%) versus Cluster 2 (**sTable 3**). Cluster 1 demonstrated higher peak serum hsTroponin I and creatinine values (**Figure 1C**). There was no difference between the clusters for biomarkers of renal inflammation or tubular injury (**Figure 1D**).

A sensitivity analysis using K-means and additional baseline biomarkers of cardiorenal disease measured only in the primary analysis cohort including NT-proBNP, NGAL, and suPAR resulted in two clinical phenotypes similar to the primary analyses (**sTable 4**). External validation, performed in 964 participants of the COPTIC cohort using K-means, also identified two clinical phenotypes similar to the primary analyses (**sTable 5**). Cluster 1, characterised by a high IL-6, iron deficiency (lower ferritin and TSat), anaemia (lower Hb) and cardiorenal dysfunction (higher Troponin, lower eGFR) phenotype experienced increased rates of death or major adverse cardiovascular events (18.3% versus 10%) (**Figure 2E),** Stage 3 acute kidney injury (15% versus 9%) and MI or new IABP post-surgery (3.7% versus 2.8%) versus Cluster 2 **(sTable 6**).

**Figure 2.**
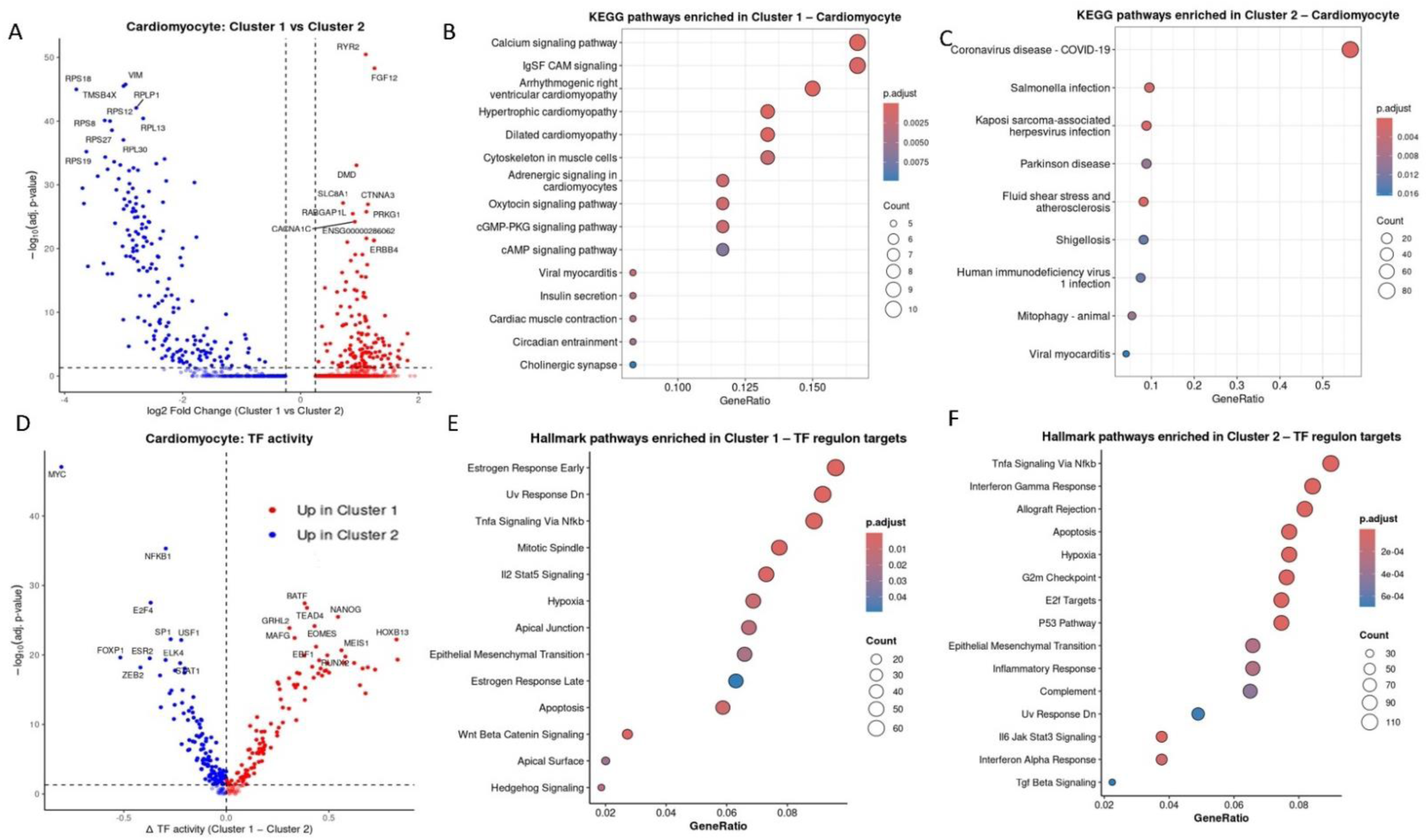
snRNAseq analysis of myocardial biopsies collected for the ObCARD study. **A**. Volcano plot showing top 20 differentially expressed genes (DEG) in cardiomyocytes from Cluster 1 versus Cluster 2. **B.** Enrichment plot showing Kyoto Encyclopaedia of Genes and Genomes (KEGG) pathway analysis of significantly DEG in Cluster 1 and **C.** Cluster 2. **D.** Volcano plot demonstrating differential TF activity in Cluster 1 versus Cluster 2. **E.** Hallmark gene set enrichment analysis of regulon targets for Cluster 1 and **F.** Cluster 2. UMAP, Uniform Manifold Approximation and Projection. TF Transcription Factor. VSMC Vascular Smooth Muscle Cell.

### Increased susceptibility to organ injury is associated with dysregulated haemopoiesis and innate immunity

In the primary analysis, Cluster 1 demonstrated attenuated leucocyte activation at baseline characterised by lower levels of activated CD64^+^/CD163^+^ leucocytes, CD14^+^/CD41^+^ monocyte-platelet aggregates, innate (CD284^+^) and adaptive (CD3^+^) immune cell-derived microvesicles and abnormalities of platelet structure and function including increases in immature platelets, reduced platelet aggregation, and increased aspirin resistance (**sTable 7**).

In the validation cohort, Cluster 1 demonstrated increased plasma levels of the immune ageing biomarkers CX3CL1/ Fractalkine, GDF-15, and CXCL9/ MIG (iAge), versus Cluster 2 (**Figure 1F**). Cluster 1 also demonstrated dysregulated haematopoiesis, characterised by lower reticulocyte counts, higher Neutrophil/Lymphocyte ratios, lower lymphocyte counts, and reduced platelet size, maturity, and platelet aggregation in response to Adrenaline at baseline, versus Cluster 2 (**Figure 1G, sTable 5**).

### snRNAseq analyses demonstrated enrichment for inflammageing and cardiomyopathy pathways in Cluster 1 and type 1 inflammation pathways in Cluster 2

We compared human atrial snRNAseq data from Cluster 1 (n=8) versus Cluster 2 (n=18) from the ObCARD subset of the primary analysis cohort (**Figure 2**). Based on the observed differences between Clusters in biomarkers of myocardial injury and innate immunity the snRNAseq analyses focused on cardiomyocytes (n=3762 nuclei) and cardiac myeloid cells (n=708 nuclei). Random down sampling was performed for the primary analyses to generate comparable numbers of nuclei in Patient Cluster 1 and Patient Cluster 2 resulting in 776 CM nuclei and 152 myeloid nuclei in each Cluster.

**For cardiomyocytes,** 319 genes showed differential expression between Cluster 1 and Cluster 2 (**Figures 2A, sTable 8**).

Cluster 1 cardiomyocytes were identified by increased expression of RYR2, FGF12, CACNA1C, and SLC8A1 and were enriched for cardiomyopathy and calcium homeostasis pathways versus Cluster 2. (**Figure 2B**, **sTables 9** and **10**). Regulon analyses identified 83 transcription factors upregulated in Cluster 1, (**Figure 2D, sTables 11** and **12**), with Hallmark pathway enrichment for stress responses; hypoxia, cGMP-PKG, oestrogen responses, dedifferentiation; Wnt, EndMT and hedgehog networks, and redox inflammation; MAPK and NF-kB networks (**Figure 2E**).

Cluster 2 demonstrated MYC driven upregulation of RPLP1, RPL13, RPS8, B2M and HLA-B genes, with pathway enrichment for cytoplasmic translation, and type 1 inflammation. (**Figure 2C, sTables 9** and **10**). Upregulated transcription factors (n=57) in Cluster 2 included FOXP1, E2F4, ELK1, and ERG, with Hallmark pathway enrichment for ribosomal biogenesis, type 1 inflammation, ribotoxic stress, and redox inflammation networks (**Figure 2F, sTables 11** and **12**).

**For cardiac myeloid cells**, sub clustering identified three subclusters; monocyte derived macrophages (MDM), tissue resident macrophages (TRM), and non-assigned macrophages (**sTable 13**).

In DEG analyses for MDM, no gene met the FDR (**sTable 14**). We therefore used PROGENY gene set enrichment analyses. This demonstrated increased TGFβ, NF-kB, and MAPK pathway signalling in Cluster 1 (**Figure 3A**). Regulon analyses demonstrated pathway enrichment for stress responses, TNFα via NF-kB, Hedgehog, and Wnt/β-catenin repair type signaling (**Figure 3 B** and **C**). MDM in Cluster 2 showed upregulation of RPL27A, S100A8 and LYZ with PROGENY demonstrating enrichment for TNFα and Hypoxia Response signaling. Regulon pathway analysis suggested enrichment for M1 type signaling (**Figures 3B** and **C, sTables 15** and **16**).

**Figure 3.**
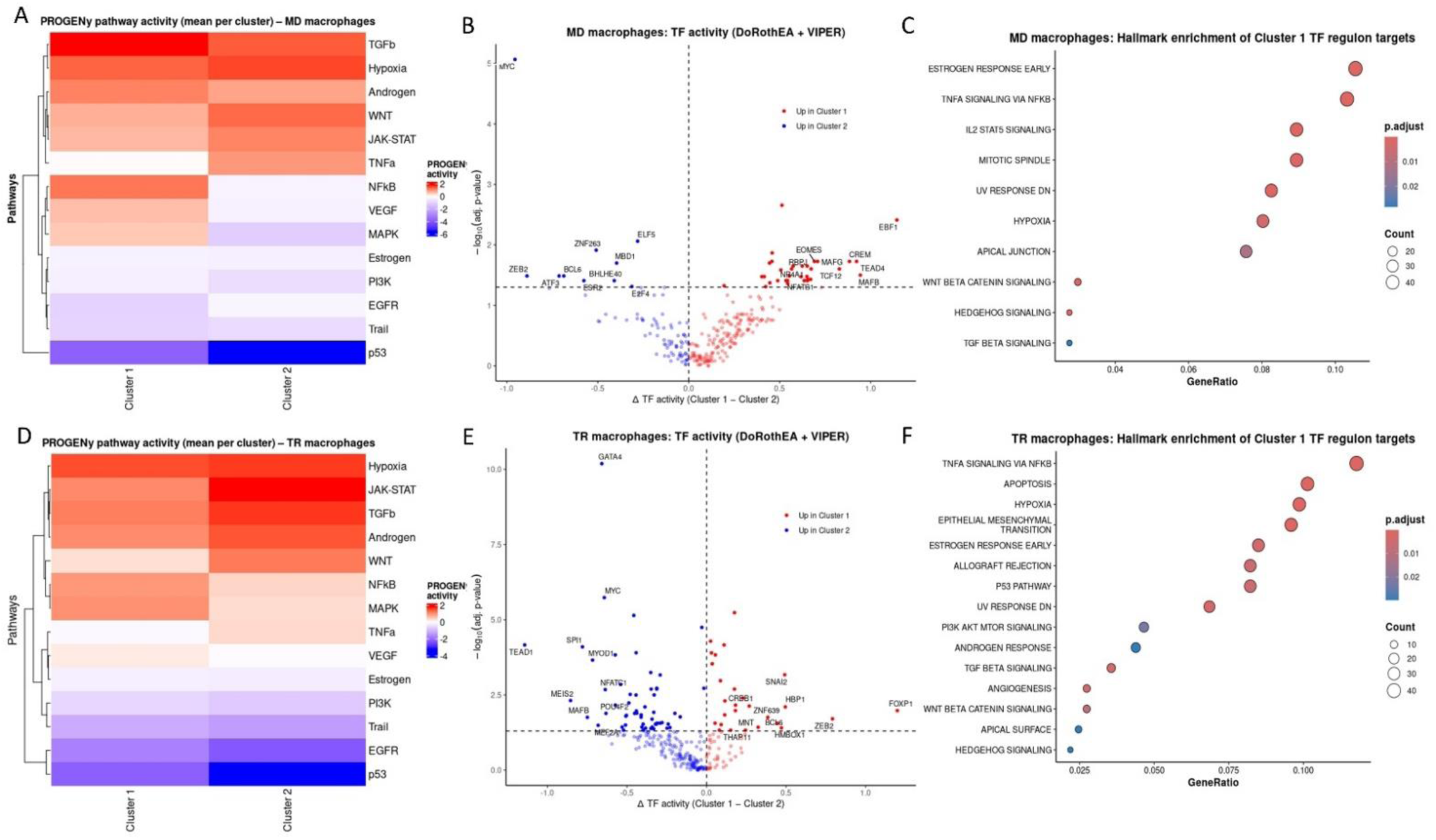
Analysis of single nuclei transcriptome in myocardial macrophages. **A.** Heatmap demonstrating PROGENY pathway enrichment for Monocyte Derived Macrophages in Cluster 1 versus Cluster 2. **B.** Volcano plot demonstrating differential TF activity in MDM from Cluster 1 versus Cluster 2. **C.** Hallmark gene set enrichment analysis of regulon targets for MDM in Cluster 1. **D.** Heatmap demonstrating PROGENY pathway enrichment for Tissue Derived Macrophages (TDM) in Cluster 1 versus Cluster 2. **E.** Volcano plot demonstrating differential TF activity in TDM from Cluster 1 versus Cluster 2. **F.** Hallmark gene set enrichment analysis of regulon targets for TDM in Cluster 1.

In TRM, Cluster 1 did not show any significantly upregulated DEG (**sTable 17**), however PROGENY pathway analyses demonstrated enrichment for anti-inflammation pathways; KLF4, FOSL1/2, RXRα. Regulon analyses demonstrated pathway enrichment for TNFα via NF-kB, epithelial–mesenchymal transition, and TGF-β signaling. In Cluster 2, DEG and pathway analyses demonstrated enrichment for cardiomyocyte genes; MYL7, MYH6, ACTC1, TNNT2, possibly indicating efferocytosis, and macrophage activation; MAFB, SPI1 (Figure 3D). Regulon analyses showed enrichment for type 1 (interferon) inflammation, mTORC1 signalling, and adaptive immune activation; IL2/STAT5 signalling (**Figure 3E** and **F, sTables 18** and **19**).

### Genetic modification of candidate genes identified from transcriptional analyses alters 90 day survival following cardiac surgery in vivo

GWAS and Mendelian Randomisation in UK Biobank showed that the genetically predicted expression of 7 genes from the cardiomyocyte snRNAseq analyses demonstrating significant differential expression between clusters with fold change>1 were causal determinants of accelerated biological ageing and 90-day mortality after surgery (**Figure 4A** and **sTable 20**). The genes; RPL31, PPP1R12B, RPL5, RPS8, RPS27A, RPS27, and SLC8A1 have functions in ribosomal translation, cellular calcium ion homeostasis, stress responses, and cardiomyocyte differentiation.

**Figure 4.**
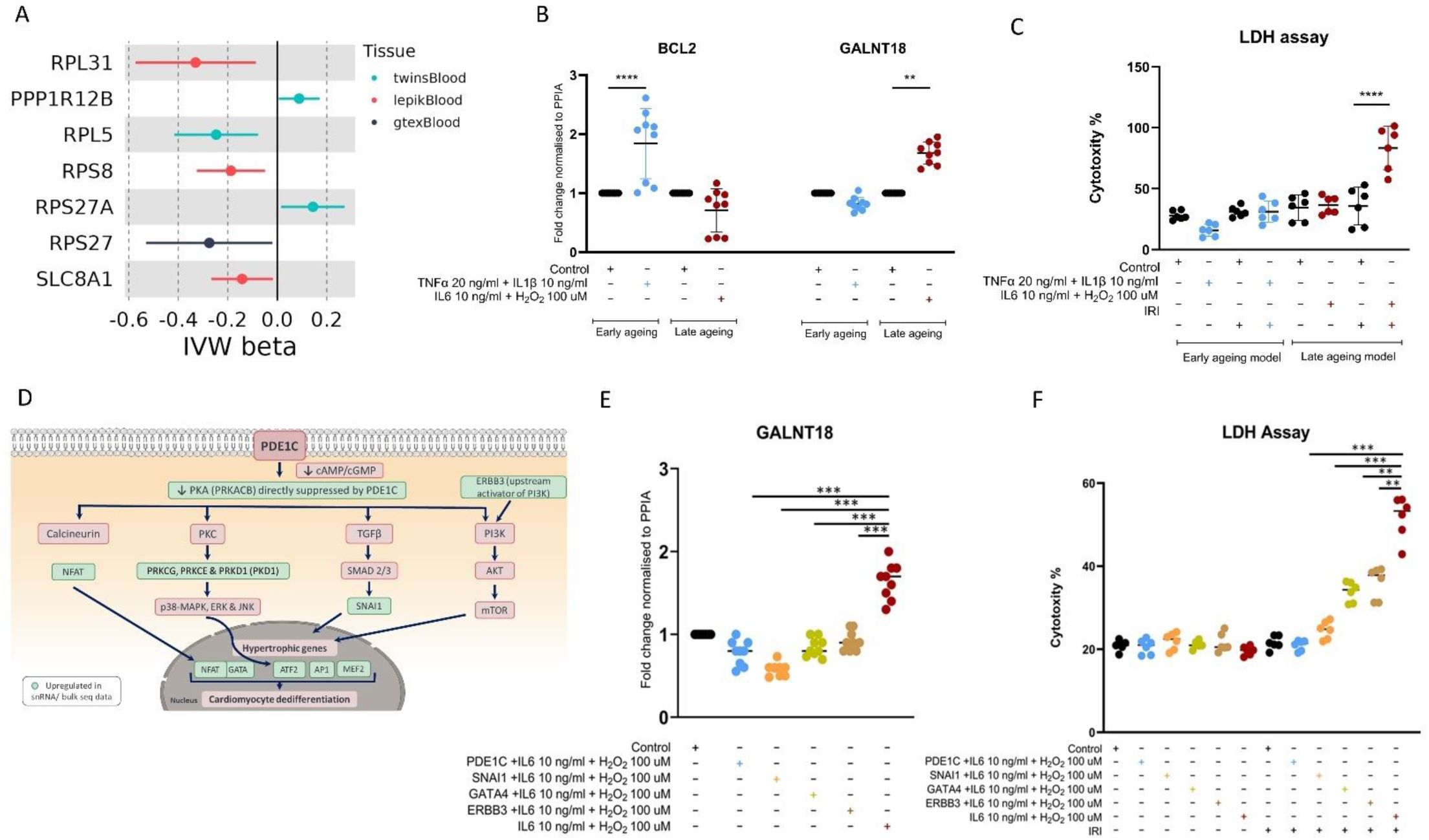
**A.** Credible causal genes from GWAS and Mendelian Randomisation in UK Biobank. **B**. Using QT-PCR, TNFα+IL-1β treated cardiomyocytes were enriched for markers of Type 1 inflammation including BCL2. IL6+H_2_0_2_treated cardiomyocytes demonstrate enrichment for markers of de-differentiation including GALNT18. **C.** IL6+H_2_0_2_treated cardiomyocytes demonstrate increased susceptibility to ischaemia reperfusion injury measured as LDH release versus TNFα+IL-1β treated cardiomyocytes. **E.** Proposed model of cardiomyocyte inflammageing in Cluster 1 based on transcriptomic data. siRNA gene knockout of key signaling nodes in cardiomyocyte inflammageing demonstrate **F.** Reductions in de-differentiation markers and **G.** Reduced susceptibility to ischaemia reperfusion injury. *P<0.05 **P<0.01 ***P<0.001 Statistical tests as per text. GWAS, Genome wide Association Study. IVW Inverse Variance Weighted. IL6, Interleukin 6. H_2_O_2_, Hydrogen Peroxide. TNFα, Tumour Necrosis factor α. GALNT18, Polypeptide N-Acetylgalactosaminyltransferase 18. BCL2, B-cell lymphoma 2. LDH, Lactate Dehydrogenase. PDE1c, Phosphodiesterase 1C. ERBB3, Erb-B2 Receptor Tyrosine Kinase 3. SNAI1, Snail Family Transcriptional Repressor 1. GATA4, GATA Binding Protein 4.

### Gene silencing of key nodes associated with the Cluster 1 Phenotype reversed cardiomyocyte susceptibility to ischaemia reperfusion injury in vitro

KEGG analysis of bulk RNAseq of cultured AC16 cardiomyocytes treated with IL6+H_2_0_2_ versus TNFα+IL-1B (**sTables 22** and **24**) demonstrated all of the differentially enriched pathways (n=416) identified from *in vivo* snRNAseq analyses of cardiomyocytes in Cluster 1 versus Cluster 2.

QT-PCR showed that cardiomyocytes treated with IL6+H_2_0_2_ expressed dedifferentiation markers (GALNT18^Hi^ BCL2^Lo^) and cardiomyocytes treated with TNFα+IL-1β expressed Type 1 inflammation markers (GALNT18^Lo^ BCL2^Hi^) (**Figure 4B**). IL6+H_2_0_2_treatment increased susceptibility to IRI versus TNFα+IL-1β **(Figure 4C**).

Transcriptomic data were used to develop a model of pathways enriched in Cluster 1 versus Cluster 2 in cardiomyocytes *in vivo* and test their role in de-differentiation and susceptibility to acute metabolic stress (**Figure 4D**). Gene silencing of key nodes associated with the Cluster 1 Phenotype using siRNA for PDE1c (cAMP/cGMP, Ca^2+^homeostasis), ERRB3 (PI3K-AKT-mTORC), SNAI1 (TGFβ-SMAD), and GATA4 (de-differentiation) reversed cardiomyocyte dedifferentiation (**Figure 4E**) and susceptibility to IRI in IL6+H_2_0_2_ treated cardiomyocytes *in vitro*, with reversal to control values of myocardial injury observed after silencing of PDE1c and SNAI (**Figure 4F**).

## Discussion

### Main findings

Unsupervised clustering analysis of people with multimorbidity (MM) undergoing cardiac surgery using pre-surgery biomarkers of haemopoietic, cardiac, metabolic, liver, and renal disease identified two distinct MM phenotypes with different levels of chronic disease progression, inflammageing, and susceptibility to organ injury.

Cluster 1 showed increased severity of cardiac disease, chronic kidney disease, iron deficiency, anaemia, higher IL-6, immune ageing, and attenuated immune cell and platelet function at baseline. Cluster 1 also developed more severe myocardial injury, acute kidney injury, major adverse cardiovascular events, and MODS post-surgery versus Cluster 2. This reflected increased susceptibility to injury given that key determinants of surgical stress; operation complexity, cardiopulmonary bypass duration, and aortic cross-clamp time were not different between clusters.

snRNAseq in Cluster 1 demonstrated enrichment for cardiomyopathy pathways and altered calcium signalling in cardiomyocytes, NF-kB, and IL-2-JAK-STAT activation in monocyte derived macrophage (MDM), and pro-fibrotic and redox inflammation signaling in tissue resident macrophages (TRM).

Genetic modification of cardiomyopathy pathways *in vivo* was associated with reductions in early mortality post cardiac surgery in UK Biobank and experimental modification of networks enriched in the Cluster 1 phenotype reduced cardiomyocyte de-differentiation and susceptibility to ischaemia reperfusion injury *in vitro*.

### Clinical Importance

First, the results are consistent with the immune system exhaustion hypothesis of inflammageing, (51) where chronic type 1 inflammation in less severe stages of chronic conditions, consistent with Cluster 2, ultimately results in inflammageing; oxidative stress mediated inflammation, myeloid exhaustion, and pro-fibrotic signalling, and chronic disease progression, consistent with the clinical characteristics of Cluster 1.

Second, the key mechanisms underlying cardiomyocyte susceptibility to injury across all of the analyses; dysregulated Ca^2+^homeostasis and proteostasis and cardiomyopathy signaling are modifiable by contemporary heart failure therapies including SGLT2 inhibitors, (52) or GLP-1 receptor agonists (53), raising the possibility that these agents may have favourable myocardial preconditioning effects if administered pre-surgery.

Third, the clusters were clinically recognisable. The injury susceptible phenotype Cluster 1 was characterised by anaemia, increased neutrophil lymphocyte ratios, altered innate immunity, and platelet dysfunction; all established risk factors for organ injury post-cardiac surgery.(54–56) Paradoxically, clinical interventions targeting individual risk factors including anaemia, platelet activation, or dysregulated innate immune responses, do not reduce organ injury.(1, 3, 57) However, the changes in Cluster 1 are also consistent with the attenuated erythropoiesis/ lymphopoiesis, and myeloid/ megakaryocyte skewing observed in bone marrow ageing.(58) In experimental studies, bone marrow ageing leads to myocardial infiltration of dysregulated bone marrow (monocyte) derived macrophages, increasing susceptibility to injury.(59–61) Our results support this hypothesis. Here, the injury susceptible phenotype was characterised by enrichment for redox-inflammation and pro-fibrosis pathways in monocyte derived macrophages associated with immune ageing, dysregulated innate immunity, and changes in myelopoiesis. The role of bone marrow ageing and monocyte derived macrophages in susceptibility to myocardial injury in people with MM merits further study.

Fourth, the study shows associations between susceptibility to myocardial injury and acute kidney injury. Acute cardiorenal injury is described in many clinical settings,(62) and the underlying mechanisms are unclear. In the current analyses, increases in acute kidney injury as determined by changes in serum creatinine were not associated with elevations in the AKI biomarkers NGAL, TIMP2*IGFBP7, and SuPAR. Recent consensus definitions of AKI incorporate this discordance between changes in eGFR and tubular injury biomarkers.(63) In previous work we have shown that therapeutic interventions targeting tubular injury do not reduce AKI.(3) Conversely, interventions that target vascular mechanisms including liberal fluid replacement or amino acid infusions prevent AKI defined by creatinine rises post-surgery.(64) Our results argue against tubular injury as a primary mechanism of cardiorenal injury post cardiac surgery even in cohorts at elevated risk.

Finally, the very high levels of post-surgery myocardial injury (63%) in Cluster 1 notwithstanding, levels of injury were also high (50%) in Cluster 2. The transcriptomic data points to different vulnerabilities in each group. For example, amplified calcium signaling; CAMK2D↑, CACNA1C↑, seen in in Cluster 1 increases the risk of Ca^2+^ overload following IRI, and subsequent cardiomyocyte necroptosis and pyroptosis.(65) In contrast enhanced ribosomal biogenesis indicated in Cluster 2 increases the risk of ribotoxic stress; RACK1↑, causing apoptosis, (66) whereas higher GPX4 expression may suppress ferroptosis. A third group of cardiac patients, without multimorbidity, also demonstrate specific myocardial signaling characteristics.(67) These results highlight the heterogeneity of baseline phenotypes, each showing different susceptibility to injury, and suggest that a precision medicine approach to organ protection will be required to deliver clinical benefits.

### Strengths and limitations

First, we used K-means clustering analyses, an unbiased machine learning method widely used for the identification of unknown clinical phenotypes. However, K-means is limited by poor reproducibility and validity. In mitigation, we demonstrated similar clinical phenotypes and outcomes when the clustering analyses used additional baseline biomarkers only available in the primary analysis cohort, and also when using biomarkers common to all the data in the external COPTIC validation cohort.

Second, the definitions of organ injury were different in the primary and validation cohorts, due to the retrospective study design. In mitigation, the directions of effect were similar for all adverse outcomes across the two clustering analysis.

Third, snRNAseq analyses were restricted to atrial tissue, and the direct applicability to ventricular susceptibility to ischaemia reperfusion injury is uncertain. We adopted this approach because the routine collection of adequate quantities of atrial myocardium is more consistent and reproducible than for ventricular tissue. In mitigation, the inflammageing injury susceptible phenotype was associated with elevated NT-proBNP, a ventricle specific marker of cardiomyocyte de-differentiation, (68) analogous to the de-differentiation pathway enrichment in atrial myocytes in the snRNAseq analyses. Our findings were consistent with recent snRNAseq analyses of accelerated biological ageing in the mouse ventricle (69) and studies linking ageing ventricular myocardium and increased susceptibility to IRI. (70) They were also validated in UK Biobank, and reproduced in a model of cardiomyocyte inflammageing and IRI *in vitro*.

### Conclusions

Inflammageing in myocardium, characterised by cardiomyopathy and redox sensitive pro-fibrotic myeloid signaling, was associated with increased levels of myocardial injury and MODS following cardiac surgery. Genetic modification of these processes *in vivo*, as well as experimental modification of these processes *in vitro* alters susceptibility to post-surgery mortality and cardiomyocyte injury respectively. The results support the evaluation of pre-surgery organ protection interventions targeting myocardial inflammageing in people referred for cardiac surgery.

## Data Availability

snRNAseq data is available at https://doi.org/10.5281/zenodo.19949025.

Anonymised clinical and biomarker data from the MARACAS, REDWASH, REVAKI-2 and COPTIC studies, as well as individual-level genetic and clinical data from the ObCARD study will be made available for ethically approved research with agreement from the studies’ sponsor.

UK Biobank data are available through a procedure described at http://www.ukbiobank.ac.uk/using-the-resource/. Annotations of UK Biobank SNPs were obtained from NealeLab data (http://www.nealelab.is/uk-biobank) round 2 results.

## Code Availability

All bioinformatic analysis were carried out in Unix, Python and R. The codes used for snRNAseq analyses are available at https://doi.org/10.5281/zenodo.19918393.

## Author Contributions

FL designed the clinical clustering analyses, acquired the data, performed the statistical analyses, and wrote the manuscript. NB and CS designed the snRNAseq analyses, acquired the UK Biobank data, and undertook the bulk of the bioinformatics analysis. SS, SL, and KT performed snRNAseq experiments and bioinformatics analyses, and helped draft the paper. MR, TC, LJD, HA, TC, AM, GA, and MZ obtained the requisite ethical approvals, performed the clinical trials, and provided the tissue and blood samples. GC and MJW coordinated and supervised laboratory analyses. GJM designed the study, wrote the manuscript, and is the guarantor of the study. All co-authors have critically reviewed the manuscript and agree to its publication.

## Protocols

Laboratory protocols for nuclei isolation in human myocardial biopsies are available at: https://doi.org/10.5281/zenodo.19921775. snRNAseq used the manufacturers protocol; Chromium Next GEM Single Cell Multiome ATAC + Gene Expression library prep: 10x genomics, CG000338 Rev F. AC16 cell line culturing also used the manufacturers protocol: Cytion, 305215.

## Data Availability

All data produced in the present study are available through approved governance processes from the University of Leicester.

## Acknowledgements

The authors are grateful to Hasmukh Patel and Julie Chamberlain at the University of Leicester who performed biomarker analyses on the COPTIC samples. We also express our gratitude to Dr Clelia Peano at Humanitas University in Milan for her expert advice and supervision on the primary analysis of the snRNAseq methods in human myocardium.

